# Real-time intelligent classification of COVID-19 and thrombosis via massive image-based analysis of platelet aggregates

**DOI:** 10.1101/2022.09.13.22279890

**Authors:** Chenqi Zhang, Maik Herbig, Yuqi Zhou, Masako Nishikawa, Mohammad Shifat-E-Rabbi, Hiroshi Kanno, Ruoxi Yang, Yuma Ibayashi, Ting-Hui Xiao, Gustavo K. Rohde, Masataka Sato, Satoshi Kodera, Masao Daimon, Yutaka Yatomi, Keisuke Goda

**Author notes:** These authors contributed equally (Chenqi Zhang, Maik Herbig). **Complete contact information:**. **Credits for research support as a footnote to the title:** N/A. **Important miscellaneous information:** N/A.

## Abstract

Microvascular thrombosis is a typical symptom of COVID-19 and shows similarities to thrombosis. Using a microfluidic imaging flow cytometer, we measured the blood of 181 COVID-19 samples and 101 non-COVID-19 thrombosis samples, resulting in a total of 6.3 million bright-field images. We trained a convolutional neural network to distinguish single platelets, platelet aggregates, and white blood cells and performed classical image analysis for each subpopulation individually. Based on derived single-cell features for each population, we trained machine learning models for classification between COVID-19 and non-COVID-19 thrombosis, resulting in a patient testing accuracy of 75%. This result indicates that platelet formation differs between COVID-19 and non-COVID-19 thrombosis. All analysis steps were optimized for efficiency and implemented in an easy-to-use plugin for the image viewer napari, allowing the entire analysis to be performed within seconds on mid-range computers, which could be used for real-time diagnosis.

## 1. INTRODUCTION

The COVID-19 pandemic has had a worldwide impact. New records in the number of global cases per day were reached in early 2022. Thrombotic complications are common in patients with COVID-19 and are the leading cause of severity (1). Studies have shown that one third of patients hospitalized for severe COVID-19 developed thrombotic complications, including venous thromboembolism, myocardial infarction, and stroke (1). In a study of a small group of COVID-19 patients in Germany, evidence of venous thromboembolism was found in 7 out of 12 COVD-19-related deaths, and massive pulmonary embolism due to lower extremity deep vein thrombosis was the direct cause of death in 4 patients (2). In addition, autopsy findings indicated that microvascular thrombosis was present in multiple organs, including the lungs, kidneys, liver, legs, heart, and brain (3), which is usually associated with multiple organ failure in severe COVID-19.

The cause of thrombosis in COVID-19 patients is not fully understood. It has been shown that venous thrombosis, arterial thrombosis, and microvascular thrombosis co-exist in COVID-19 (1). Additionally, coagulopathy, complement activation, cytokine release, platelet hyperactivity, thrombocytopathy, and vascular endothelial dysfunction are potentially major factors in the pathogenesis of thrombosis in COVID-19 patients (4). Although COVID-19-associated thrombosis appears to have similar mechanisms to non-COVID-19 thrombosis, the disease progresses much more rapidly, which could herald a distinction between COVID-19 and non-COVID-19 thrombosis.

An imaging flow cytometry (IFC) technique, based on frequency-division-multiplexed (FDM) microscopy that can capture blur-free bright-field images of fast-flowing cells, has previously been used in studies comparing the size of platelet aggregation as the initiation of microthrombus formation (5). Their results have shown an increased concentration of platelet aggregates in COVID-19 patients. This technology has been used in the study of the effects of COVID-19 vaccines on platelets, showing that standard doses of the Pfizer-BioNTech (BNT162b2) vaccine have negligible effects on platelets (6). This technique can be used to study the morphology of platelet aggregates and has the potential to reveal morphological differences between COVID-19 and non-COVID-19 thrombosis, which may be of significant help in the detection of COVID-19 and in the development of antithrombotic treatment strategies for COVID-19 patients (7).

In this paper, to better understand the difference between COVID-19 and non-COVID-19 thrombosis, we report a machine learning approach to discriminate between blood samples from patients with COVID-19 and non-COVID-19 thrombosis by large-scale single-cell image-based analysis and by interpreting trained machine learning models. As shown in Figure 1, our analysis approach consists of (i) high-throughput bright-field imaging of platelets (including single platelets and platelet aggregates) and white blood cells (WBCs) in the blood of patients with COVID-19 and non-COVID-19 thrombosis by IFC based on FDM microscopy (see Figure S1 and the Materials and Methods section for details), (ii) convolutional neural networks (CNNs) to distinguish between single platelets, platelet aggregates, and WBCs, (iii) a random forest (RF) model (8) and a cumulative distribution transform (CDT)-based classification algorithm (9) for classifying diseases based on the morphological features of the cells and aggregates. As a results, we identified blood samples from patients with COVID-19 and non-COVID-19 thrombosis using the machine learning approach we developed, with a test accuracy of 75%. In the morphological analysis of platelets, both the RF and CDT approach show that the platelet size distribution tends to widen in patients with COVID-19. This wider platelet size distribution indicates an alteration in platelet production or clearance (10). Finally, we implemented the CNN, RF, and CDT-based analysis methods in an easy-to-use napari plug-in (https://www.napari-hub.org/plugins/disease-classifier) which was integrated into the workflow directly after IFC measurements. Using this plugin, measurements typically containing 25,000 images can be processed in less than 6 seconds, which can be applied for real-time classification after measurement.

**FIGURE 1.**
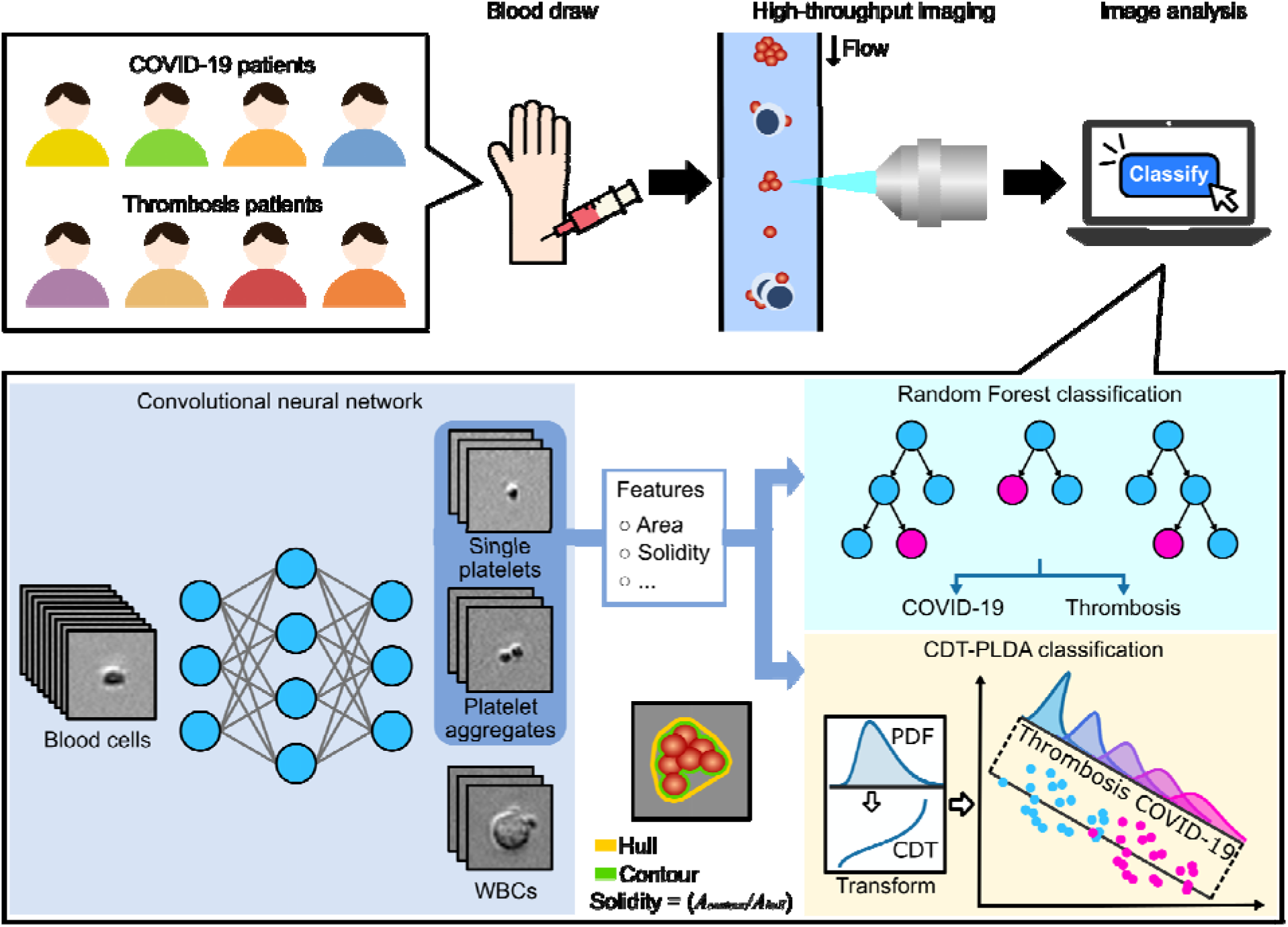
Conceptual schematic of the intelligent classification workflow, including sample preparation, high-throughput IFC measurement, CNN-based phenotype classification, computation of morphological features (area and solidity), and PLDA-based disease classification.

## 2. MATERIALS AND METHODS

### 2.1 Human subjects

We enrolled data from patients who were hospitalized at the University of Tokyo Hospital between November 2020 and August 2021 and diagnosed with COVID-19 or thrombosis. 101 samples for thrombosis and 181 samples for COVID-19 were obtained from 41 thrombosis (54% male, 46% female) and 34 COVID-19 (79% male, 21% female) patients. The initial laboratory test values and characterization for each patient are listed in Table S1. COVID-19 was diagnosed using reverse transcription polymerase chain reaction (RT-PCR). All the COVID-19 patients received mechanical ventilation or extracorporeal membrane oxygenation. Thrombosis was diagnosed using ultrasonography or computed tomography and RT-PCR was used to verify that all thrombosis patients were COVID-19 negative. After clinical laboratory tests, residual coagulation test samples (with 3.2% citrate) were collected from the patients (in accordance with the ethical approvals no. 11049 and no. 11344, granted by the Institutional Ethics Committee in the School of Medicine at the University of Tokyo). Clinical data and laboratory tests were retrieved from the electronic medical patient records using a standardized data collection protocol. Patients gave informed consent to participate in the study and had the option to opt-out via the webpage of the University of Tokyo Hospital. Patients who opted out were excluded from our study. 500 µL of blood was diluted in 5 ml of saline (0.9% NaCl). After adding the sample to Lymphoprep solution (STEMCELLS, ST07851), a density gradient centrifugation protocol (800 g for 20 min) was performed according to a protocol provided by the vendor to isolate the platelets. After centrifugation, 500 µL were taken from the mononuclear layer, and platelets were stained using 10 µL of anti-CD61-PE (Beckman Coulter, IM3605) and 5□µL of anti-CD45-PC7 (Beckman Coulter, IM3548). To conserve platelet aggregates, the sample was fixed by adding 500□µL of 2% paraformaldehyde (Wako, 163-20145). The blood cell suspension was inserted into the microchannel of the FDM-based IFC for measurement.

### 2.2 FDM microscope

FDM microscopy is an imaging method that was used to capture blur-free bright-field images of cells moving at a high speed of 1 m/s, lending itself well to IFC (11). Figure S1 shows the schematic of the IFC system that consists of a microfluidic chip, a syringe pump, and an FDM microscope. The cells were hydrodynamically focused to the center of a microfluidic channel where they were imaged by the FDM microscope. The FDM microscope consists of a continuous-wave (CW) laser (491 nm, Cobolt Calypso, 491□nm, 100□mW), beam splitters, and acousto-optic deflectors (Brimrose TED-150-100-488, 100-MHz bandwidth), generating a spatially distributed frequency comb which was used as the light source for bright-field imaging. A single-pixel photodetector (Thorlabs APD430A/M) was employed for imaging, which was triggered based on the CD61-PE fluorescence signal. The captured waveforms were reconstructed into bright-field images using LabVIEW (LabVIEW 2016), resulting in 67 × 67 pixel images with a spatial resolution of 0.8 μm/pixel. For each measurement, 25,000 images were acquired at an event rate of 100 – 300 events per second (eps). An efficient contour detection algorithm was implemented using OpenCV (12) (see Supplementary Information for details). From the contour, the cross-sectional objective area (A) and solidity (S) were determined (Figure 1).

### 2.3 CNN model for phenotype classification

Images containing noise, platelets, platelet aggregates, and WBCs were loaded into AIDeveloper (v. 0.2.3), a software program for training deep neural networks (13). Random sampling was employed to balance the dataset. To prevent the model from overfitting, we used the following image augmentation operations: rotation, flip, horizontal or vertical shift, brightness change, Gaussian noise, and Gaussian blur. We selected a CNN with four convolutional layers and a total number of 475,362 trainable parameters. During training, AIDeveloper (13) saved models automatically when a new record in validation accuracy or validation loss was reached.

### 2.4 RF model for disease classification

RFs are a class of supervised machine learning models based on an assembly of decision trees that are trained on random subsets of the dataset (14). For classification, the majority vote of all trees was used. Decision trees were trained to find thresholds to separate the data into classes. The ability of one specific feature to split the data into classes were quantified by the Gini impurity index, also called feature importance. In this work, we used the Python package scikit-learn (v. 1.0.2) to train RF models and obtain the feature importance. We employed a weighting of the loss function to equalize the contribution of each class of the unbalanced dataset of this study.

### 2.5 Cumulative distribution transform (CDT) for disease classification

We considered 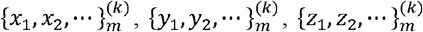 and 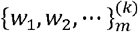 to be the sets of the morphological feature measurements corresponding to the n^th^ subject of the k^th^ disease class (thrombosis or COVID-19). Here, x, y, z, and w denote the area of platelets, the solidity of platelets, the area of platelet aggregates, and the solidity of platelet aggregates, respectively. Also, we considered 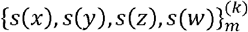 to be a set of univariate probability density functions (PDFs) obtained from the morphological feature measurements using a kernel density estimation technique. The goal of the classification problem was to determine the class of a test set, {*s*(*x*), *s*(*y*), *s*(*z*), *s*(*w*)} corresponding to a subject with an unknown diagnosis. The first step was to obtain the transformed versions of the PDFs, denoted as 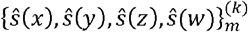 using the cumulative distribution transform (CDT) (15). The CDT is a map from the space of smooth PDFs to the space of diffeomorphisms, which can be defined as the inverse function of the cumulation of each individual PDF. The CDT enhances linear separability in data by removing certain nonlinearities and simplifies the classification problem (15). The CDT is an invertible, one-to-one, and differentiable map, which enabled us to interpret the trained model by visualizing the class differences obtained by the model (Figure 1). After CDT transformation, we employed principal component analysis to reduce data dimensionality using scikit-learn (v. 1.0.2). For classification, we employed the penalized linear discriminant analysis (PLDA) classifier (16), which differentiated between the classes of a given dataset by obtaining the most discriminant directions computed based on Fisher’s linear discriminant in combination with penalized least-squares regression. We used the Python package PyTransKit (v. 0.2.3) to compute the CDTs and train the PLDA classifier and the Python package statsmodels (v. 0.13.2) to obtain the PDFs using the kernel density estimation technique.

## 3. RESULTS

### 3.1 Data acquisition

101 samples for non-COVID-19 thrombosis and 181 samples for COVID-19 were measured using our FDM-based IFC. For each sample, image acquisition was triggered until 25,000 CD61+ events were captured. An efficient image segmentation algorithm was implemented using OpenCV to determine the projected area and solidity of each event. Figure 2A shows a scatter plot of the events in area vs. solidity of one representative measurement. Each captured image may contain noise, a single platelet, or a WBC. Representative example images for these phenotypes are shown as figure insets in Figure 2A. The scatter plot does not show separate populations for platelets, platelet aggregates, and WBCs because their area and solidity distributions are overlapping. Hence, for a meaningful analysis, it is essential to discriminate between these subpopulations.

**FIGURE 2.**
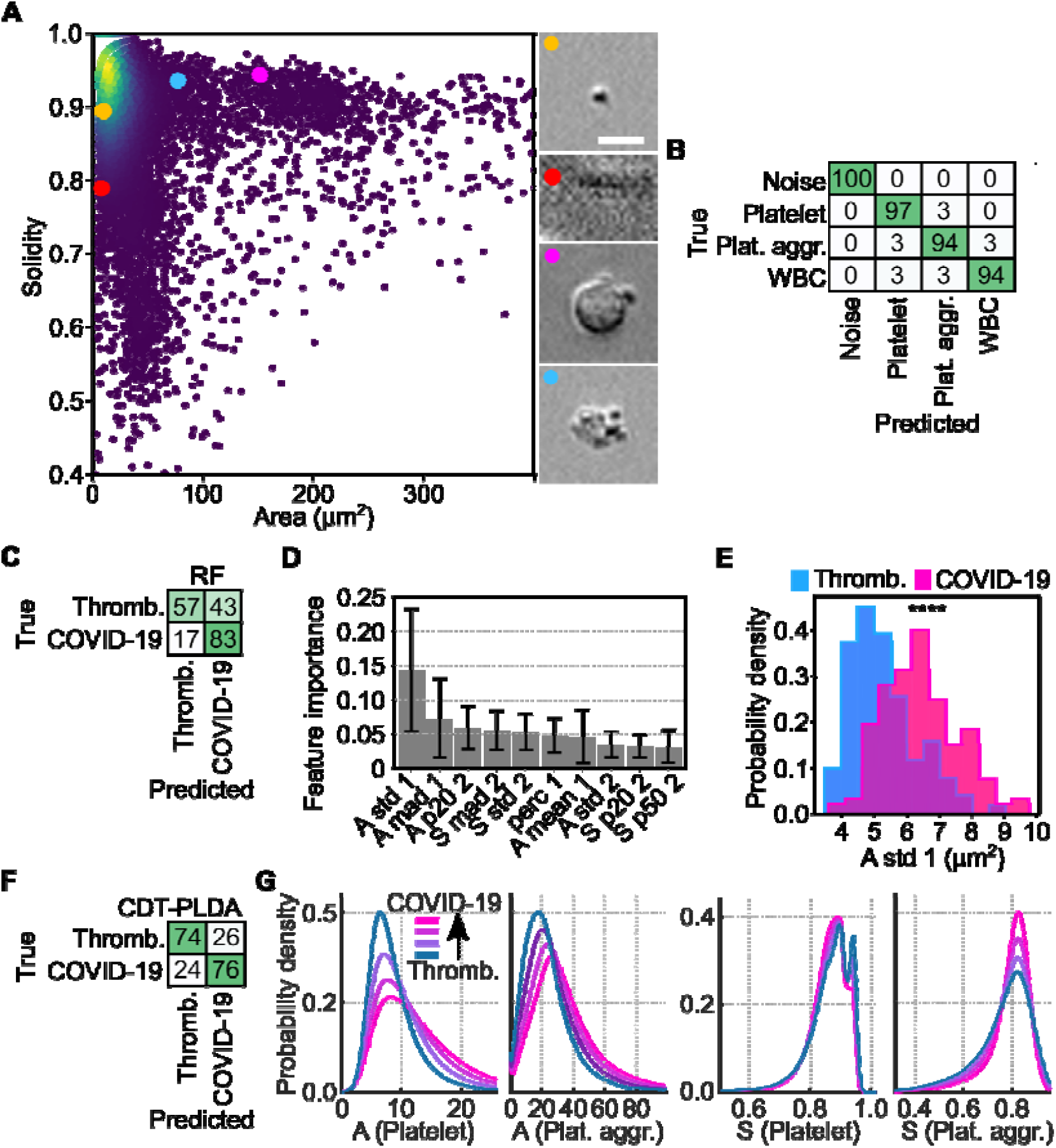
Machine learning methods for differentiating COVID-19 and non-COVID-19 thrombosis. (A) Scatter plot that shows a representative measurement of non-COVID-19 thrombosis. Inset images show example phenotypes of noise, platelets, platelet aggregates, and WBCs. Colored dots show corresponding locations of those events in the scatter plot. Scale bar = 10 μm. (B) Normalized confusion matrix that shows the performance of the CNN for phenotype classification. (C) Normalized confusion matrix (in %) that shows the disease classification performance of RF models on the testing dataset (average of 1000 iterations). (D) Bar plot that shows the mean and std of the feature importance of the 10 most important features, resulting from 1000 RF models that were trained using different measurements for the training dataset. (A -area, S – solidity, std – standard deviation, mad – median absolute deviation, 1 – platelet, 2 – aggregate) (E) Histogram that shows the distribution of the feature with the highest feature importance. Stars indicate statistical significance, determined via a two-sided t-test (****: p < 10^−4^). (F) Normalized confusion matrix that shows the classification performance of PLDA models on the testing dataset (average of 1000 iterations). (G) Histograms that show reconstructed distribution profiles, corresponding to different locations along the most significant CDT-PLDA direction. Blue and magenta lines show distributions that resemble more thrombosis-like and more COVID-19-like distribution shapes, respectively.

### 3.2 CNN-based phenotype classification

Using YouLabel (v. 0.2.4), we manually labeled 37199, 66611, 12269, and 10274 images of noise, platelets, platelet aggregates, and WBCs, respectively. We employed AIDeveloper to load all the images and train a CNN with four convolutional layers to discriminate these phenotypes, resulting in a validation accuracy of 96% (see the developed confusion matrix in Figure 2B and Figure S2). Due to the low complexity of the CNN, it only took 2.8 s (on an Intel Core i5-8257U CPU at 1.40GHz, 8GB RAM) to obtain the predictions of a whole measurement with approximately 25,000 events. The trained CNN was applied to all measurements of COVID-19 and non-COVID-19 thrombosis. The predictions were employed to split the dataset. We computed the mean, standard deviation (std), median absolute deviation (mad), the 20^th^, 50^th^, and 80^th^ percentile (p20, p50, p80) for the area (A) and solidity (S) of platelets and platelet aggregates, individually. These features describe the shape of a distribution in terms of location and width. Furthermore, the percentage of the events of single platelets and platelet aggregates was computed, resulting in a total of 26 features that describe each measurement.

### 3.3 RF model for disease classification

To identify which feature or combination of features allows COVID-19 to be distinguished from non-COVID-19 thrombosis, we trained a supervised machine learning model. The model development process was carried out in 3 successive steps. First, we selected 10 random measurements of COVID-19 and non-COVID-19 thrombosis to create a validation set and 10 further random measurements of both diseases to create a testing dataset. Second, the remaining 81 non-COVID-19 thrombosis and 161 COVID-19 measurements were employed to train a RF model for classification. To optimize the number of trees (n_trees_) in the model, we trained RF models having 1 ≤ n_trees_ ≤ 100 trees and chose the model with maximum validation accuracy. Third, this model was evaluated by computing the testing accuracy. Moreover, we obtained the feature importance values (see the Materials and Methods section). A high accuracy could occur by chance, depending on which measurements were used for training, validation, and testing. Therefore, we repeated steps one, two, and three 1000 times, resulting in a distribution of testing accuracies and feature importance values. The testing accuracy was found to be (mean ± std) 69.9% ± 9.3%. Figure 2C shows the normalized confusion matrix for the testing data, averaged over 1000 iterations. The distribution of feature importance values of the 10 most important features is displayed in Figure 2D (see Figure S3A for all features), showing that the most important features describe the width of the distribution of platelet sizes (A_std_1_, A_mad_1_), the 20^th^ percentile of the area of platelet aggregates (A_p20_2_), and the width of the distribution of solidity of platelet aggregates (S_mad_2_, S_std_2_). Interestingly, the width of the distribution of platelet sizes A_std_1_ (shown in Figure 2E) is significantly larger (p = 1.8×10^−15^) for COVID-19 compared to thrombosis, meaning that the distribution of shape features can be used to discriminate the two from each other. However, the selection of distribution shape parameters (such as standard deviation) was arbitrary and is unlikely optimal. Therefore, below we discuss a method for automatic determination of distribution-shape features, based on the PDF of the distribution.

### 3.4 CDT-based model for disease classification

To obtain the PDF, we performed a kernel density estimation using the feature values (A, S). The distributions were sampled over a uniform grid of N = 5,000 points. Next, the PDF was transformed using the CDT, which returned a feature vector of length N. To reduce data dimensionality, we used principal component analysis and selected the principal components such that the sum of variance explained was 99% of the total variance. For training the PLDA classification model, we employed the same iterative approach as for the RF model training. The model reached an average testing accuracy of 75.3% ± 9.1%. The normalized confusion matrix in Figure 2F shows the average performance of the CDT-PLDA of 1000 iterations. To interpret the CDT-PLDA model, we used one model that was trained on the entire dataset and plotted the distribution profiles along the most significant direction, as shown in Figure 2G. Blue and magenta curves indicate distribution shapes that are typical of non-COVID-19 thrombosis and COVID-19, respectively.

### 3.5 napari plugin for real-time inference

We implemented the phenotype prediction (using the CNN) and disease classification (using the RF and PLDA) into a plugin for napari with an intuitive and interactive user interface (Figure S4, Video S1). Napari is a Python-based image viewer, specialized for large datasets, which is easy to install. Our plugin allows users to load a measurement file (typically 25,000 images) by drag and drop and perform the analysis pipeline outlined in this paper in less than 6 s (on an Intel Core i5-8257U CPU at 1.40GHz, 8GB RAM). The interactive user interface of the plugin allows the selection of phenotype classes and displays corresponding images. Using efficient OpenCV implementations, we achieved a computational time of 2.82 s per measurement for performing background subtraction, cell segmentation, and phenotype classification using the CNN. The subsequent disease classification using the RF or PLDA model only required 0.03 s, or 1.6 s, respectively. The plugin is an open-source software program, available on the napari-hub (https://www.napari-hub.org/plugins/iacs-ipac-reader), which can be installed and applied without the need for programming knowledge.

## 4. DISCUSSION

In this paper, we report a proof-of-concept application of an FDM-based IFC system for analyzing platelets in COVID-19 and non-COVID-19 thrombosis. We trained a CNN to identify single platelets and platelet aggregates in measurements of patient blood. After deriving morphological features, we trained RF- and PLDA-based classification models to distinguish COVID-19 and non-COVID-19 thrombosis, reaching an average testing accuracy of 69.9% and 75.3%, respectively. Moreover, we identified morphological features that were significantly different between COVID-19 and non-COVID-19 thrombosis. We implemented the workflow into a plugin for napari, allowing the analysis to be performed in less than 6 s without the need for programming. These results indicate that the combination of IFC, machine and deep learning, and napari could be employed to study platelet aggregate formation and would even allow for real-time diagnostics.

The accurate discrimination of subpopulations required care. Image acquisition was triggered for CD61+ events (platelets) and the scatter plot in Figure 2A shows a broad range of cell sizes and various phenotypes (platelets, platelet aggregates, WBCs, see inset images in Figure 2A). Unspecific staining, for example, due to the presence of debris from platelets, could cause the trigger signal for WBCs. Therefore, the discrimination of subpopulations was performed before subsequent analysis, which was achieved by training a CNN based on manually labelled images. The CNN for phenotype classification showed a high accuracy of 96%. In fact, for all wrongly classified cells (see the confusion matrix in Figure 2B and Figure S2), even manual labeling was challenging.

After the phenotype classification, we computed area and solidity for platelets and platelet aggregates separately. Based on the distribution-shape of area and solidity, we trained the RF and PLDA models for the classification of COVID-19 and non-COVID-19 thrombosis. The RF model received manually defined distribution features such as the mean and std as input. In contrast, for the CDT-PLDA, we used the CDT and PCA to compute and select distribution features. The confusion matrices in Figure 2C and Figure 2F show that the latter approach is superior, indicating that the selected distribution features for the RF missed important distribution-shape characteristics. The leftmost histogram in Figure 2G, where curves are non-overlapping, indicates that the area of platelets was different between COVID-19 and non-COVID-19 thrombosis. While the location of the peak is similar for non-COVID-19 thrombosis (blue) and COIVD-19 (magenta), the tail of the distribution is much more pronounced for COVID-19, resulting in a wider distribution. This finding agrees with the result of the RF, which found high importance for A_std_1_.

Our analyses show that the distribution of platelet size is wider in COVID-19 (Figure 2E, Figure 2G), implicating that larger thrombocytes are more abundant. This is consistent with the previous reports that platelets in COVID-19 are highly reactive and have a unique transcriptome profile, and that platelet size and maturity are associated with increased critical illness and all-cause mortality in hospitalized COVID-19 patients (17). Larger platelets are generally at higher risk of forming aggregates and promoting thrombus formation (18). Although there are reports that cardiovascular outcomes are associated with platelet size (19), platelet size was more pronounced in COVID-19 in our analysis. Moreover, the rightmost histogram in Figure S3B shows that the distribution of the solidity values of platelet aggregates is significantly lower for COVID-19 (p = 3.5×10^−10^), which means that irregularly shaped platelet aggregates occur more often in the blood of COVID-19 patients. The irregular morphology of platelet aggregates may reflect differences in the mechanisms underlying their formation. (20)

To make the analysis methods shown in this paper easily accessible, we developed a user-friendly plugin for napari. It allows users to perform the disease classification between COVID-19 and non-COVID-19 thrombosis in less than 6 s and results can be seen in a single glance. The plugin promotes the translation of basic science to clinical application as it could be employed for real-time diagnostics. The plugin is not limited to the CNN, RF, and CDT-PLDA models shown in this work, but also allows the loading of models for different or complementary analyses.

This paper shows a proof-of-concept demonstration that could be extended in various directions. For analysis, we only employed events of platelets and platelet aggregates. However, we also observed platelets attached to WBCs, which could correspond to platelet-monocyte aggregates whose occurrence is related to platelet activation (18). In a future study, additional staining would allow interpretation of these events, such that including them in the overall analysis could be valuable. Furthermore, we limited the morphology quantification to cell size and solidity. A more extensive quantification of cell shape and texture could allow improvement of classification accuracy. Moreover, the CDT-PLDA analysis indicated that further distribution shape features such as skew and kurtosis would be promising features for prospective improvement of the RF model. For phenotype classification, we employed a relatively small CNN architecture to promote execution speed and applicability on hardware with low or medium performance. Using a GPU, larger models could be run in real time which may further lift the accuracy. The current experimental setup requires an image reconstruction step after the measurement. Parallelizing signal acquisition and image reconstruction would not only improve CPU utilization but would also lead to a more seamless real-time experience. In addition, upgrading the current imaging system by introducing the capability of capturing more high-content information, such as quantitative optical phase, may further enrich our image analysis and increase testing accuracy. Finally, we included both arterial thrombosis (e.g., cerebral and myocardial infarction) and venous thrombosis (e.g., deep vein thrombosis and pulmonary embolism) in the non-COVID-19 thrombosis patient group. While similarities between them have been reported in recent years, they have different pathological mechanisms, which may suggest that their platelet aggregates have different morphological features (21). To address this point, a further study is needed to increase the number of patients with a variety of thrombosis and compare their images with those in patients with COVID-19 using our method.

## Data Availability

All raw data and Python scripts for analysis are available on Zenodo: https://doi.org/10.5281/zenodo.6825004. The napari plugin is available on the napari-hub (https://www.napari-hub.org/plugins/disease-classifier).

## ACKNOWLEDGEMENTS

This work was supported mainly by AMED JP20wm0325021 (M. N., K. G., Y. Y.) and JSPS Core-to-Core Program (K. G.) and partly by JSPS KAKENHI grant numbers 19H05633, 20H00317 (K. G.), and 21K15640 (M. N.), White Rock Foundation (K. G.), Ogasawara Foundation (K. G.), Nakatani Foundation (K. G.), Konica Minolta Foundation (K. G.), National Institutes of Health R01 GM130825 (G. K. R., MSER), National Science Foundation 1759802 (G. K. R., M. S. E. R.), and Charitable Trust Laboratory Medicine Research Foundation of Japan (M. N.). C. Z. was supported by Epson International Scholarship.

## AUTHOR CONTRIBUTIONS

Chenqi Zhang: Formal analysis; investigation; methodology; software; visualization; writing-review & editing. Maik Herbig: Data curation; formal analysis; methodology; project administration; software; supervision; writing-review & editing. Yuqi Zhou: Data curation; investigation; writing-review & editing. Masako Nishikawa: Data curation; investigation; writing-review & editing. Mohammad Shifat-E-Rabbi: Formal analysis; methodology; software; visualization; writing-review & editing. Hiroshi Kanno: Data curation. Ruoxi Yang: Data curation. Yuma Ibayashi: Data curation. Ting-Hui Xiao: Methodology; project administration; resources; supervision; software; writing-review & editing. Gustavo K. Rohde: Methodology; supervision; writing-review & editing. Yutaka Yatomi: Methodology; project administration; resources; supervision; writing-review & editing. Keisuke Goda: Funding acquisition; project administration; resources; supervision; writing-review & editing.

## CONFLICT OF INTEREST

K. G. is a shareholder of two cell analysis startups (CYBO and Cupido). All other authors declare no competing interests.

## S1 Image segmentation

## Supporting Information

Since cells are hydrodynamically focused to the center of the channel, the border of the image was employed to obtain the mean background brightness, which was subtracted from each image. Since cells can be brighter or darker than the background, we computed the absolute of the pixel values after subtraction of the background brightness. To identify a threshold for distinguishing cells from the background, we computed the 95th percentile of the background pixel intensities using all images of the dataset. After binarization of images using the threshold, a contour finding algorithm was applied (22). The projected cell area was obtained from the contour. After obtaining the convex hull of the contour, the solidity was computed as the ratio of the areas of contour and hull.

**FIGURE S1.**
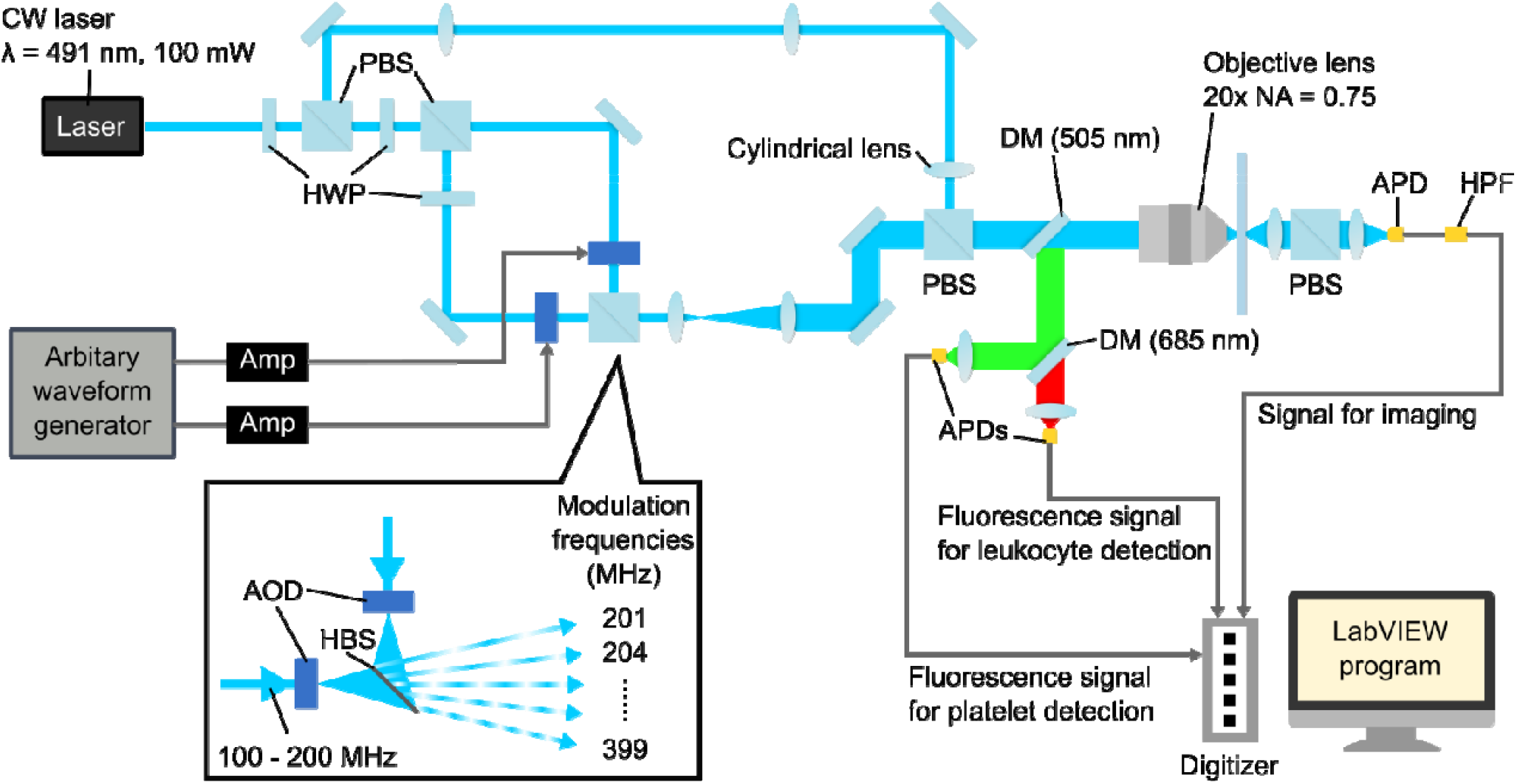
Detailed sketch of the FDM-based imaging setup. CW: continuous wave; PBS: polarizing beam splitter; HWP: half-wave plate; DM: dichroic mirror; NA: numerical aperture; Amp: Amplifier; APD: avalanche photodetector; HPF: high-pass filter; HBS: half beam splitter; AOD: acousto-optic deflector.

**TABLE S1.** Demographics, clinical characteristics, and laboratory findings of patients. All the patients in this study were hospitalized at the University of Tokyo Hospital. Data are expressed as median values (IQR), n (%), or n/N (%). p values were calculated by the Mann-Whitney U test or Fisher’s exact test.

|  | All patients | Thrombosis patients | COVID severe patients | p value |
| --- | --- | --- | --- | --- |
|  | n = 75 | n = 41 | n = 34 |  |
| Age, years | 71 (60.5-77.5) | 74.0 (64.0-80.0) | 67.5 (59.0-72.0) | 0.0095 |
| Sex |  |  |  | 0.0281 |
| Male | 49 (65%) | 22 (54%) | 27 (79%) |  |
| Female | 26 (35%) | 19 (46%) | 7 (21%) |  |
| Comorbidity |  |  |  |  |
| Hypertension | 40 (53%) | 22 (54%) | 18 (53%) | 1.0000 |
| Diabetes | 21 (28%) | 10 (24%) | 11 (32%) | 0.6061 |
| Coronary heart disease | 5 (7%) | 3 (7%) | 2 (6%) | 1.0000 |
| Chronic kidney disease | 9 (12%) | 6 (15%) | 3 (9%) | 0.4989 |
| Obesity, BMI > 25 | 21 (28%) | 11 (27%) | 10 (30%) | 1.0000 |
| Active malignancy | 13 (17%) | 11 (27%) | 2 (6%) | 0.0293 |
| Diagnosis of thrombosis | 45 (60%) | 41 (100%) | 3 (9%) | <0.0001 |
| Thrombosis site |  |  |  |  |
| Deep vein thrombosis | 16 (21%) | 16 (39%) | 0 (0%) | <0.0001 |
| Pulmonary embolism | 6 (8%) | 6 (15%) | 0 (0%) | 0.0290 |
| Cerebral infarction | 11 (15%) | 10 (24%) | 1 (3%) | 0.0096 |
| Myocardial infarction | 7 (9%) | 7 (17%) | 0 (0%) | 0.0140 |
| Others | 4 (5%) | 2 (5%) | 2 (6%) | 1.0000 |
| Antithrombotic therapy at baseline | 25 (33%) | 19 (46%) | 6 (18%) | 0.0133 |
| Antithrombotic therapy during treatment | 64 (85%) | 30 (73%) | 34 (100%) | 0.0007 |
| Antiplatelet therapy at baseline | 20 (27%) | 15 (37%) | 5 (15%) | <0.0001 |
| Anticoagulation therapy at baseline | 9 (12%) | 7 (17%) | 2 (6%) | 0.3263 |
| Antiplatelet therapy during treatment | 12 (16%) | 12 (29%) | 0 (0%) | 0.0003 |
| Anticoagulation therapy during treatment | 56 (75%) | 22 (54%) | 34 (100%) | <0.0001 |
| Laboratory findings |  |  |  |  |
| Leukocyte count, $\times 10^9/L$ | 7.9 (5.7-11.8) | 6.9 (5.2-8.0) | 10.9 (7.7-13.3) | 0.0003 |
| Red cell count, $\times 10^{12}/L$ | 384.0 (323.0-437.0) | 357.0 (307.0-409.0) | 405.5 (371.0-450.0) | 0.0086 |
| Platelet count, $\times 10^9/L$ | 20.7 (14.1-28.1) | 21.2 (11.8-29.6) | 20.6 (16.7-27.7) | 0.4531 |
| ALT, U/L | 36.5 (19.0-76.0) | 22.0 (13.0-52.0) | 58.0 (32.0-132.0) | 0.0003 |
| Creatinine, $\mu\text{mol}/L$ | 0.8 (0.7-1.3) | 0.7 (0.6-1.5) | 0.9 (0.7-1.2) | 0.5722 |
| Lactate dehydrogenase, U/L | 350.5 (231.0-560.0) | 245.0 (205.0-328.0) | 529.5 (381.5-683.0) | <0.0001 |
| C-reactive protein, mg/L | 4.2 (1.2-8.6) | 2.1 (0.4-7.1) | 5.3 (2.0-14.0) | 0.0156 |
| PT-INR | 1.1 (1.0-1.2) | 1.1 (1.0-1.2) | 1.1 (1.0-1.2) | 0.5053 |
| APTT, s | 29.9 (26.3-25.7) | 28.2 (25.2-34.9) | 32.1 (27.6-40.0) | 0.0566 |
| D-dimer, $\mu\text{g}/\text{mL}$ | 9.6 (2.3-23.1) | 14.6 (4.4-32.6) | 4.6 (2.0-13.1) | 0.0280 |

**FIGURE S2.**
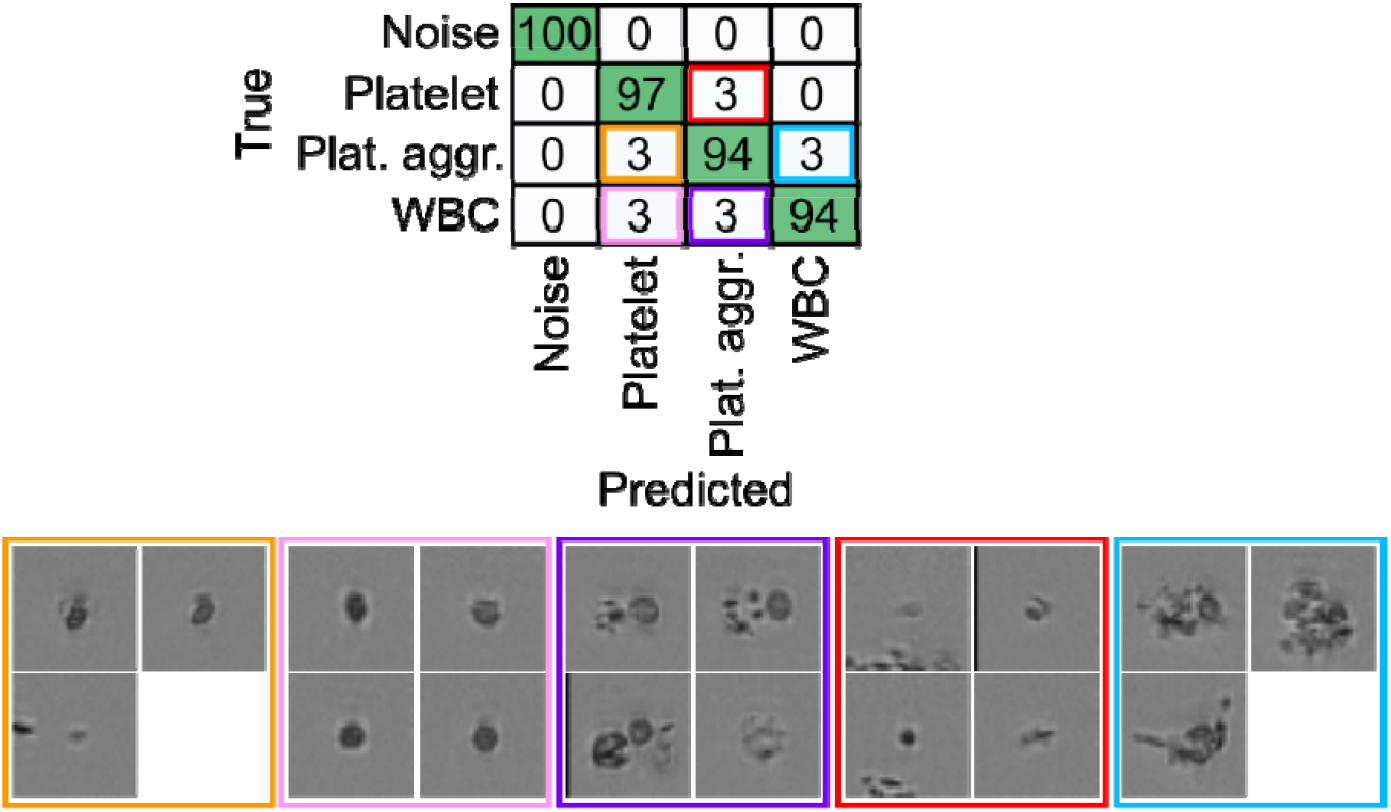
Normalized confusion matrix that shows the result of the CNN for phenotype classification being applied to the validation set. Non-diagonal elements correspond to classification errors which are highlighted in colors, and example images are provided for each. In each case, the decision of the CNN seems reasonable and even manual labeling would be challenging. Hence, avoiding this type of model error would require more differentiated ground truth information for example by previously enriching cells, or by further staining of cell types.

**FIGURE S3.**
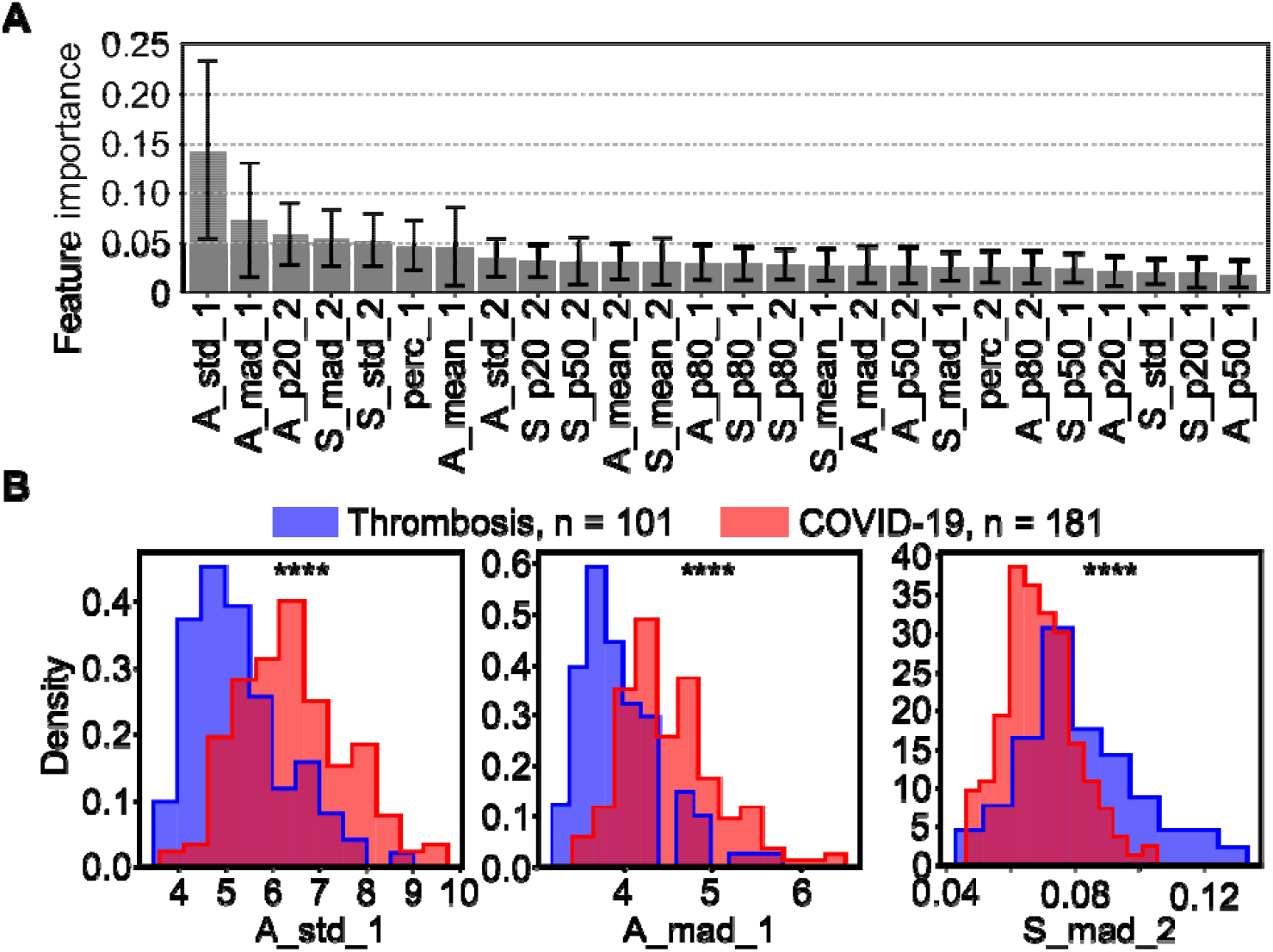
Details of the RF analysis. (A) Bar plot that shows the mean and std of the feature importance of the 10 most important features, resulting from 1000 RF models that were trained using different measurements for the training dataset. (B) Histogram that shows the distribution of the three features with the highest feature importance. The width of the distribution of platelet sizes (A_std_1, A_mad_1), is significantly larger (p = 1.8·10^−15^ and p = 1.5·10^−14^) for COVID-19 compared to non-COVID-19 thrombosis. Moreover, the width of the distribution of platelet aggregate’s solidity (S_mad_2), is significantly smaller (p = 3.5·10^−10^) for COVID-19 compared to non-COVID-19 thrombosis.

**FIGURE S4.**
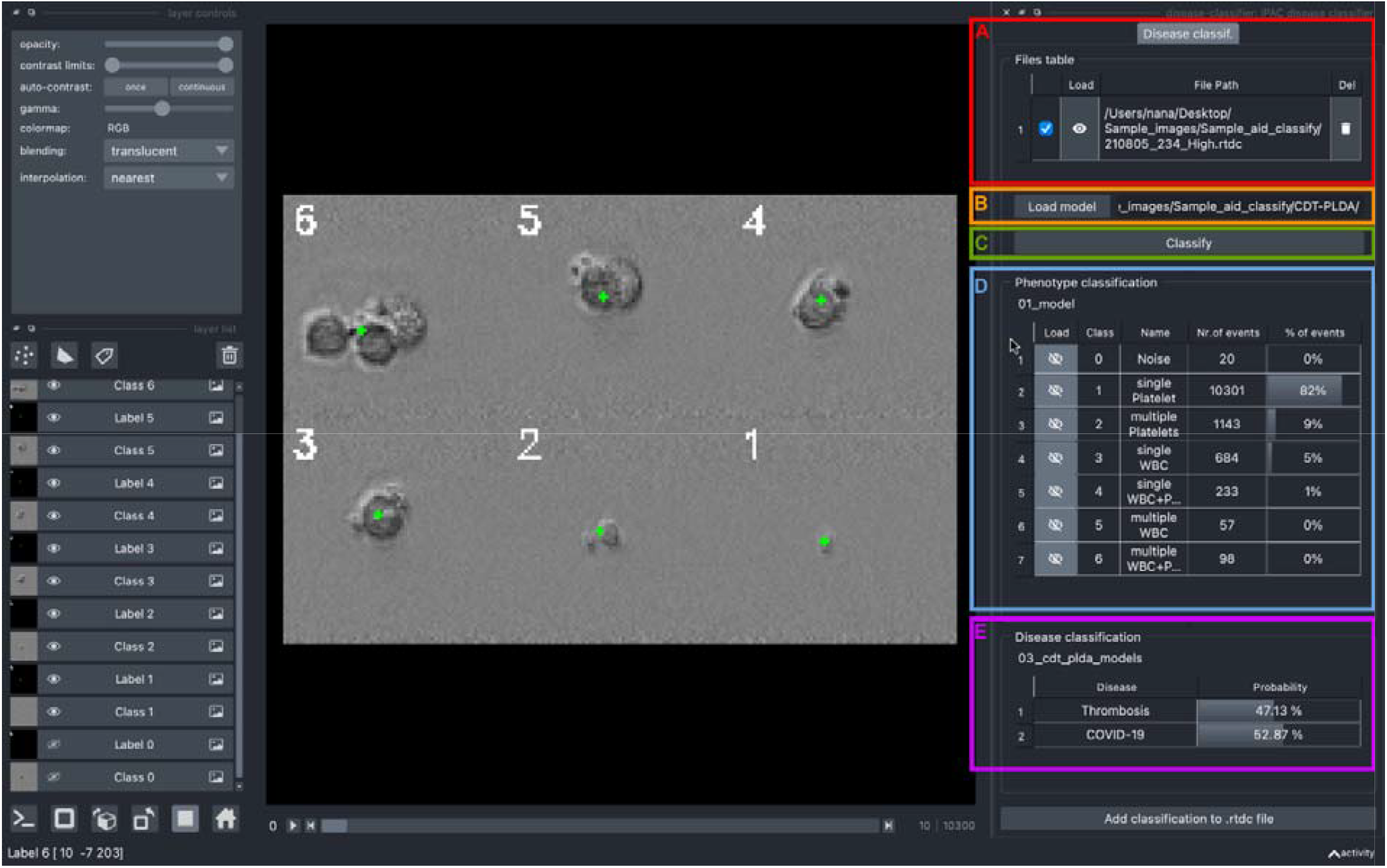
Graphical user interface of napari and the napari plugin. (A) The region indicated by the red rectangle allows users to load measurement files by drag-and-drop. (B) The GUI element for loading the CNN for phenotype classification and RF or CDT-PLDA model for disease classification. Besides the models shown in the present manuscript, alternative models could also be loaded. (C) The GUI element used to start the analysis. Subsequently, the results are displayed below. (D) The Blue rectangle indicates the region where phenotype classification results are displayed. The interactive table allows users to display images of a class by clicking the icon in the leftmost column. Cell images are shown in the napari viewer (center) (E) Magenta region shows the disease classification resulting from the RF or CDT-PLDA model. The columns show the disease types and corresponding probabilities.

## Notes

### Author Declarations

ethical approvals no. 11049 and no. 11344, granted by the Institutional Ethics Committee in the School of Medicine at the University of Tokyo

